# Altered Spatiotemporal Dynamics of Self-Referential Processing in Bipolar Disorder

**DOI:** 10.64898/2026.08.30.26361790

**Authors:** Pao-Huan Chen, Niall W. Duncan, Yi-Ju Liu, Hsin-Chien Lee, Tzu-Yu Hsu

## Abstract

**Background:** Bipolar disorder is associated with persistent social, cognitive, and functional impairment during euthymia, yet the neural mechanisms underlying these deficits remain unclear. Alterations to self-referential processing are a candidate mechanism, but existing electrophysiological studies rely on emotionally valenced paradigms that potentially confound self-processing with emotional biases.

**Methods:** We analysed electroencephalography from 28 patients with bipolar disorder (type I or II) and 28 age- and sex-matched healthy controls during an emotionally neutral colour judgment task with self-related (preference) and non-self-related (similarity) conditions. Late positive potentials, temporal generalisation decoding, and frequency band decoding (theta, alpha, beta) were used to characterise the temporal dynamics and oscillatory correlates of self versus non-self processing.

**Results:** Controls showed higher overall event-related potential amplitudes and greater self versus non-self differentiation than patients (condition by group interaction, 337 to 946 ms). Broadband temporal generalisation decoding revealed extensive cross-temporal generalisation of the self versus non-self representation in controls, spanning most of the trial, but no significant generalisation in patients. Frequency analyses showed that alpha and beta carried self versus non-self information in both groups, with broader extent in controls, and that anterior theta carried this information in patients but not controls. Exploratory correlations linked decoding measures to rumination and anxiety but not to manic symptoms.

**Conclusions:** The neural representation distinguishing self-referential from externally guided processing was both smaller in amplitude and less temporally sustained in bipolar disorder. Reduced persistence is not detectable by conventional amplitude analyses, and may bear on the self-related and social cognitive difficulties reported in this population.

## Introduction

Bipolar disorder is associated with persistent functional impairment that extends well beyond acute mood episodes. Even during euthymia, patients show deficits in social cognition, mentalizing, and self-other processing that predict disability and reduced quality of life (Lewandowski et al., 2024; Sevindik et al., 2025). These impairments cannot be fully explained by residual mood symptoms or medication effects, suggesting that a core neurocognitive deficit, or set of deficits, underlies the functional difficulties observed during clinical remission (Martínez-Arán et al., 2004).

The nature of any such deficit(s) remains an outstanding question, with different alternatives having been proposed (Keramatian et al., 2021). One potential candidate is an alteration in self-related processing in bipolar disorder. This refers to processes involved in the evaluation of external stimuli in relation to oneself, deficits in which could influence multiple downstream processes. From a social cognition perspective, alterations to how one perceives the self and its separation from others could affect an understanding of how others think and their relation to oneself (Lasagna et al., 2026; Lewandowski et al., 2024). At the same time, processes such as working memory (Yin et al., 2019, 2021) and metacognition (Vaccaro & Fleming, 2018) have been described as being linked to alterations in self-related processing in general, with some evidence from bipolar disorder in particular (Porta-Casteràs et al., 2023).

Self-related processing has been linked to cortical midline structures, including the medial prefrontal cortex and posterior cingulate cortex, which overlap with the default mode network (Northoff et al., 2006; Qin & Northoff, 2011). These regions show altered functional properties across phases of bipolar disorder (Pomarol-Clotet et al., 2015), and disrupted self-continuity and identity disturbance are recognized as core clinical features of the condition (Inder et al., 2008). Self-referential processing also generates distinct electrophysiological signatures, particularly late positive potentials over frontocentral regions, that index sustained evaluative engagement (Auerbach et al., 2015, 2016), and is linked to theta band oscillatory activity at midline sites (Knyazev, 2013). Despite the potential clinical relevance of self-referential processing to functional recovery in bipolar disorder, the temporal dynamics and oscillatory mechanisms of this cognitive function remain poorly understood in patients who are clinically stable and no longer in acute mood episodes.

Only a handful of studies have directly examined the neural basis of self-referential processing in bipolar disorder, and none have used paradigms that isolate self-processing from emotional content. An event-related potential study using a self-referential memory task found that larger P300 amplitudes for self-referential relative to other-referential conditions were present in controls but absent in bipolar patients (Zhao et al., 2016), and a subsequent analysis of the same paradigm reported weaker response power and phase lag index values in patients, with group differences confined to the theta and alpha bands (Zhang et al., 2022). Disrupted switching between the default mode and central executive networks during a self-referential functional magnetic resonance imaging task has been observed in euthymic bipolar patients (Porta-Casteràs et al., 2023), and altered resting state electroencephalographic microstates in euthymic patients may reflect abnormal self-focused processing (Vellante et al., 2020). A recent systematic review of neural oscillations in bipolar disorder confirmed that abnormal oscillatory activity,

particularly in the alpha and beta bands, is a consistent finding across mood states (Su et al., 2024), building upon foundational comparative work demonstrating pervasive resting-state EEG power and coherence abnormalities in the disorder (Kam et al., 2013). All existing electroencephalographic studies of self-referential processing in bipolar disorder, however, have employed either resting-state conditions or paradigms involving emotionally valenced stimuli, making it impossible to separate self-referential processing from emotional biases. In addition, no study has applied multivariate temporal decoding methods to characterize the representational dynamics of self versus non-self processing in bipolar disorder, despite growing recognition that such methods can reveal cognitive processing abnormalities invisible to univariate approaches in psychiatric populations (Marsicano et al., 2024).

A previous study from our group addressed the emotional valence confounded by employing an emotionally neutral colour judgment paradigm to investigate self-referential processing in major depressive disorder (Hsu et al., 2021). The task involves a preference condition representing self-related processing and a similarity condition representing non-self-related processing, using identical visual stimuli across conditions (Johnson et al., 2005), and converging EEG and fMRI evidence has confirmed that these two conditions engage dissociable neural networks including default mode and perceptual regions (Ratnasari et al., 2025). Patients with major depressive disorder showed a sustained difference in late positive potential amplitudes between self and non-self conditions at frontocentral electrodes, absent in controls. This difference correlated positively with depressive symptoms and rumination. Elevated theta oscillatory power at central electrodes during the self-related condition was also specific to the patient group. These findings demonstrated that altered self-referential processing in depression can be isolated from emotional content biases inherent in trait attribution tasks (Benau et al., 2019; Herbert et al., 2011). The emotionally neutral nature of this paradigm makes it particularly suitable for bipolar disorder, where emotional dysregulation could otherwise obscure self-specific processing differences.

The present study applied this paradigm to bipolar disorder with two methodological advances. First, temporal generalisation decoding (King et al., 2014; King & Dehaene, 2014) was used to characterize whether neural representations of self versus non-self processing are sustained, transient, or sequentially transformed over time. Unlike traditional univariate event-related potential analyses, this multivariate approach captures fine-grained, distributed neural patterns and has recently been used to successfully decode responses to emotionally salient self-referential statements in depression and suicidality (Jeong et al., 2026). We hypothesised that bipolar patients would show reduced late positive potential differentiation between conditions and reduced temporal stability of the self versus non-self neural representation compared to controls.

Second, we further characterised the oscillatory and spatial organisation of this processing. Temporal generalisation decoding was performed separately on theta, alpha, and beta band power (Grootswagers et al., 2017; Li et al., 2024; Xie et al., 2020), and separately for anterior and posterior electrode subsets, motivated by the frontocentral topography of the late positive potential and the association between frontal midline theta and self-referential processing (Hsu et al., 2021; Knyazev, 2013). Theta was examined because of its established link to self-referential processing, alpha because of its role in attentional gating (Klimesch, 2012) and its documented reduction in euthymic bipolar disorder (Başar et al., 2012), and beta because of its involvement in evaluative maintenance (Engel & Fries, 2010) and its reported abnormalities in bipolar disorder (Su et al., 2024). We further examined whether the resulting decoding measures were associated with residual depressive and anxiety symptoms and ruminative tendencies. These analyses were exploratory and intended to generate hypotheses about the oscillatory mechanisms underlying any observed differences in temporal stability.

## Methods and Materials

### Participants

Thirty-two patients with bipolar disorder (type I or II; 24 female; mean age 31.2 ± 8.1 years) were recruited from the Department of Psychiatry at Taipei Medical University Hospital and diagnosed by experienced psychiatrists according to DSM-5. All were on a stable medication schedule for at least four weeks and did not meet criteria for a current manic, hypomanic, or depressive episode at testing (BPRS 25.8 ± 6.0; YMRS 2.0 ± 4.5; HAMD-17 6.0 ± 4.7; HAMA 6.8 ± 5.4). Thirty-four age- and sex-matched healthy controls (25 female; mean age 31.9 ± 7.2 years) with no psychiatric or neurological history and no psychotropic medication use were recruited from the community. Groups did not differ in age (t(62) = 0.66, p = 0.511) or sex distribution (Fisher’s exact p = 1.000). Ruminative tendencies were assessed with the 22-item Ruminative Response Scale (controls 38.0 ± 10.1; patients 52.9 ± 16.7). After exclusion of two patients without colour task data and eight participants with fewer than 20 valid trials per condition, electroencephalographic analyses included 28 controls and 28 patients. The study was approved by the Joint Institutional Review Board of Taipei Medical University (N202106045, N202204047), and all participants gave written informed consent. Further detail is given in Supplementary Methods and Supplementary Tables 1 and 2.

### Colour judgment task

Three coloured squares were presented in a triangular arrangement, one target above two comparison squares, using colours drawn from a set of 12 equiluminant values in CIELAB space. In the similarity condition the comparison squares differed in distance from the target and participants judged which was closer in hue; in the preference condition both comparisons were equidistant from the target and participants chose the pairing they preferred. Converging electroencephalographic and functional magnetic resonance imaging evidence indicates that the two conditions engage dissociable networks including default mode and perceptual regions (Ratnasari et al., 2025) (Ratnasari et al., 2025). Each trial comprised a blank screen (500 to 1000 ms), a fixation cross (1000 ms), the stimulus array (2000 ms), and a response prompt (1500 ms); responses were not accepted before the prompt appeared. Conditions were presented in separate counterbalanced blocks of 150 trials each. Stimulus parameters, and the modifications relative to Hsu et al. (2021), are given in Supplementary Methods.

### Electroencephalographic recording and analysis

Electroencephalograms were recorded from 30 electrodes (Easycap; BrainAmp) at 1000 Hz. Following independent component analysis for ocular and muscular artifacts, data were filtered at 0.1 to 45 Hz. Event-related potential epochs (−200 to 2500 ms) were baseline-corrected to the prestimulus interval; epochs for frequency analyses (−1000 to 2500 ms) were left uncorrected in the time domain and normalised in the frequency domain. Epochs exceeding ±100 µV were rejected, trials without a response and incorrect similarity trials were excluded, and trial counts were equalised across conditions within participants.

Group, condition, and interaction effects on event-related potentials were tested with spatiotemporal cluster-based permutation tests (5000 permutations). Temporal generalisation decoding (King & Dehaene, 2014) was performed in MNE-Python (Gramfort et al., 2013) using linear discriminant analysis with 5-fold stratified cross-validation, training a classifier at each time point and testing it at all others. Because the event-related potential analysis established a condition by group interaction, decoding was run within each group separately: on broadband data, on theta (5 to 7 Hz), alpha (8 to 13 Hz), and beta (14 to 31 Hz) power derived from Morlet wavelets (Xie et al., 2020), and separately for anterior and posterior electrode subsets. Matrices were tested against chance with two-dimensional cluster-based permutation tests (5000 permutations). Per-participant mean AUC within significant anterior clusters was correlated with HAMD-17, HAMA, and RRS scores using Spearman correlations. Full parameters are given in Supplementary Methods.

## Results

### Behavioral results

Behavioural analyses were conducted on a sample of 64 participants (34 HC, 30 BD), as two patients were excluded prior to analysis due to missing response. In the similarity condition, accuracy was high in both groups but was higher in controls than in patients (controls: 0.93 ± 0.04; patients: 0.88 ± 0.10; t = 2.78, p = 0.007, Cohen’s d = 0.68; Figure 1b left panel). For reaction times, a 2 (condition: similarity, preference) × 2 (group: controls, patients) mixed ANOVA revealed a main effect of group (F(1,62) = 4.76, p = 0.033, ηp² = 0.07; Figure 1b middle panel), with patients responding more slowly overall. A main effect of condition was also observed (F(1,62) = 5.51, p = 0.022, ηp² = 0.08). A condition × group interaction (F(1,62) = 4.73, p = 0.034, ηp² = 0.07) indicated that group differences in reaction time were more pronounced for the similarity condition (controls: 410 ± 103 ms; patients: 494 ± 159 ms; t = −2.53, p = 0.014) than for the preference condition (controls: 408 ± 100 ms; patients: 458 ± 139 ms; t = −1.67, p = 0.099). Although there were these statistical group differences, accuracy remained high in both groups (>87%) and no-response rates were low (<2%; Figure 1b right panel), suggesting that all participants engaged meaningfully with the task.

**Figure 1.**
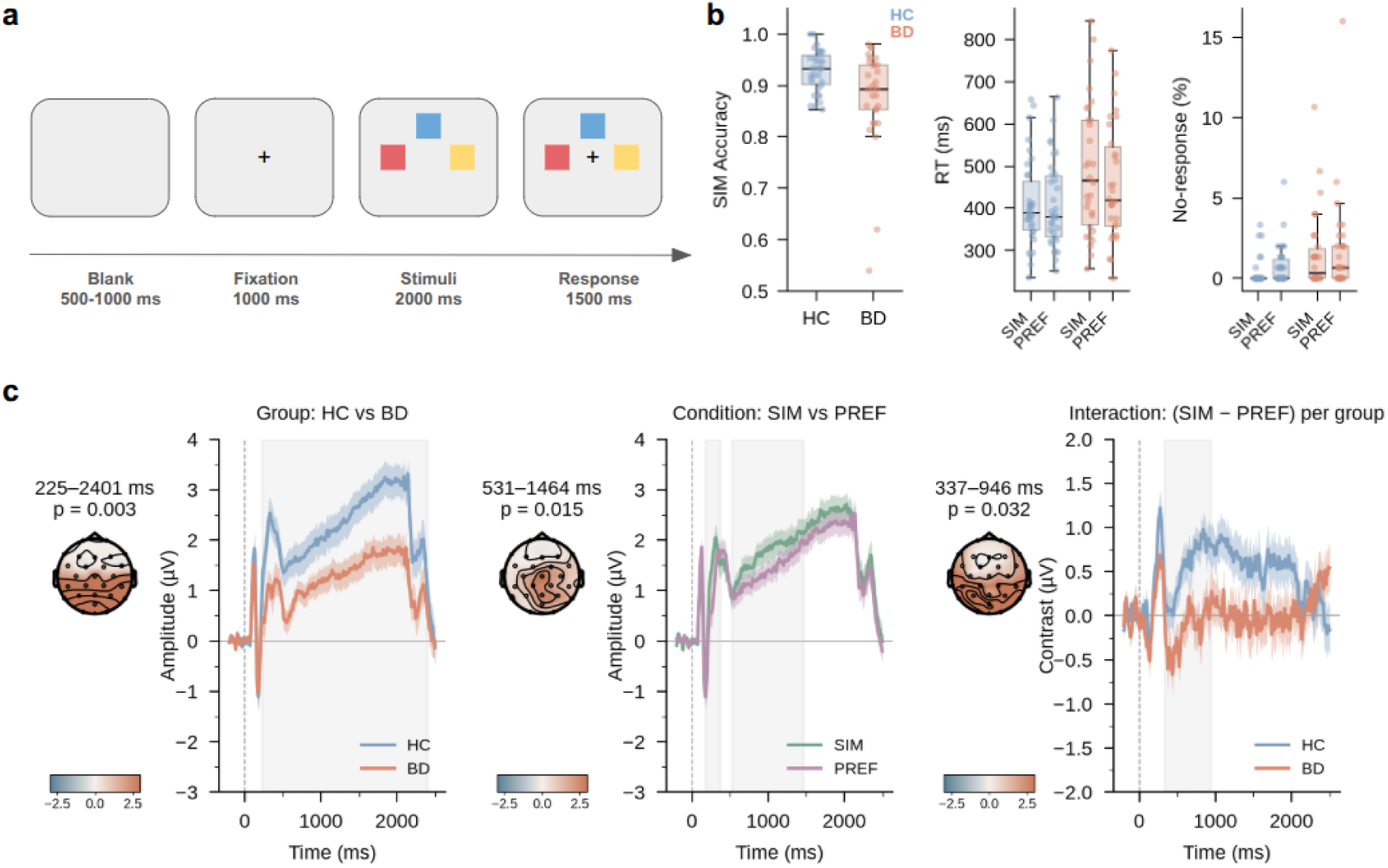
Task design, behavioral performance, and event-related potential results. (a) Experimental procedure. Each trial began with a blank screen (500 to 1000 ms), a fixation cross (1000 ms), a stimulus array of three coloured squares (2000 ms), and a response prompt (1500 ms response window). In the similarity condition, the two comparison squares differed in distance from the target (30° and 60°) and participants judged which was most similar. In the preference condition, both comparison squares were equidistant (60°) and participants chose which pairing they preferred. Conditions were presented in separate blocks (150 trials each). (b) Behavioral performance. Left: accuracy in the similarity condition for healthy controls (HC; n = 34) and patients with bipolar disorder (BD; n = 30). Right: reaction times for each condition and group. BD showed lower accuracy and a significant condition by group interaction for reaction times. Error bars indicate standard error of the mean. (c) Event-related potential results. Grand-averaged waveforms and topographic maps showing three significant effects from spatiotemporal cluster-based permutation tests. A group main effect (225 to 2401 ms) indicated higher amplitudes in HC than BD. Two condition clusters reflected greater amplitudes for similarity at centro-parietal sites (181 to 375 ms and 531 to 1464 ms). A condition by group interaction (337 to 946 ms) confirmed greater self versus non-self differentiation in HC than BD. SIM indicates similarity condition and PREF indicates preference condition.

### Event-related potentials

Spatiotemporal cluster-based permutation tests on event-related potential amplitudes (n = 28 per group) revealed three effects (Figure 1c). A main effect of group emerged within a cluster spanning 225 to 2401 ms across 18 channels (cluster p = 0.003, Figure 1c left panel), indicating that controls exhibited overall higher amplitudes than patients throughout the sustained processing window. A main effect of condition was observed in two positive clusters: an early cluster from 181 to 375 ms across 23 channels (cluster p = 0.044) and a later cluster from 531 to 1464 ms across 22 channels (cluster p = 0.015; Figure 1c middle panel), both indicating greater amplitudes for the similarity condition at centro-parietal and occipital sites. A condition by group interaction was seen within a cluster spanning 337 to 946 ms across 25 channels (cluster p = 0.032; Figure 1c right panel), confirming that the magnitude of differentiation between self and non-self conditions was greater in controls than in patients. There was weak statistical evidence for a second interaction cluster from 962 to 1476 ms across 19 channels (p = 0.052).

### Whole-brain temporal generalisation decoding

To investigate the temporal stability and dynamics of the neural representations underlying task performance, we applied whole-brain temporal generalisation decoding. Broadband temporal generalisation decoding (n = 28 per group) revealed a significant condition effect (p < 0.01; Figure 2b left panel). Controls showed extensive temporal generalisation (p < 0.001, training 105 to 2435 ms, testing 95 to 2435 ms; Figure 2b right panel), with a broad, square-shaped cluster indicating a sustained and stable neural code maintained throughout the trial (King and Dehaene, 2014). Patients showed no significant temporal generalisation (Figure 2b middle panel). The direct group comparison did not reach significance (Figure 2a).

**Figure 2.**
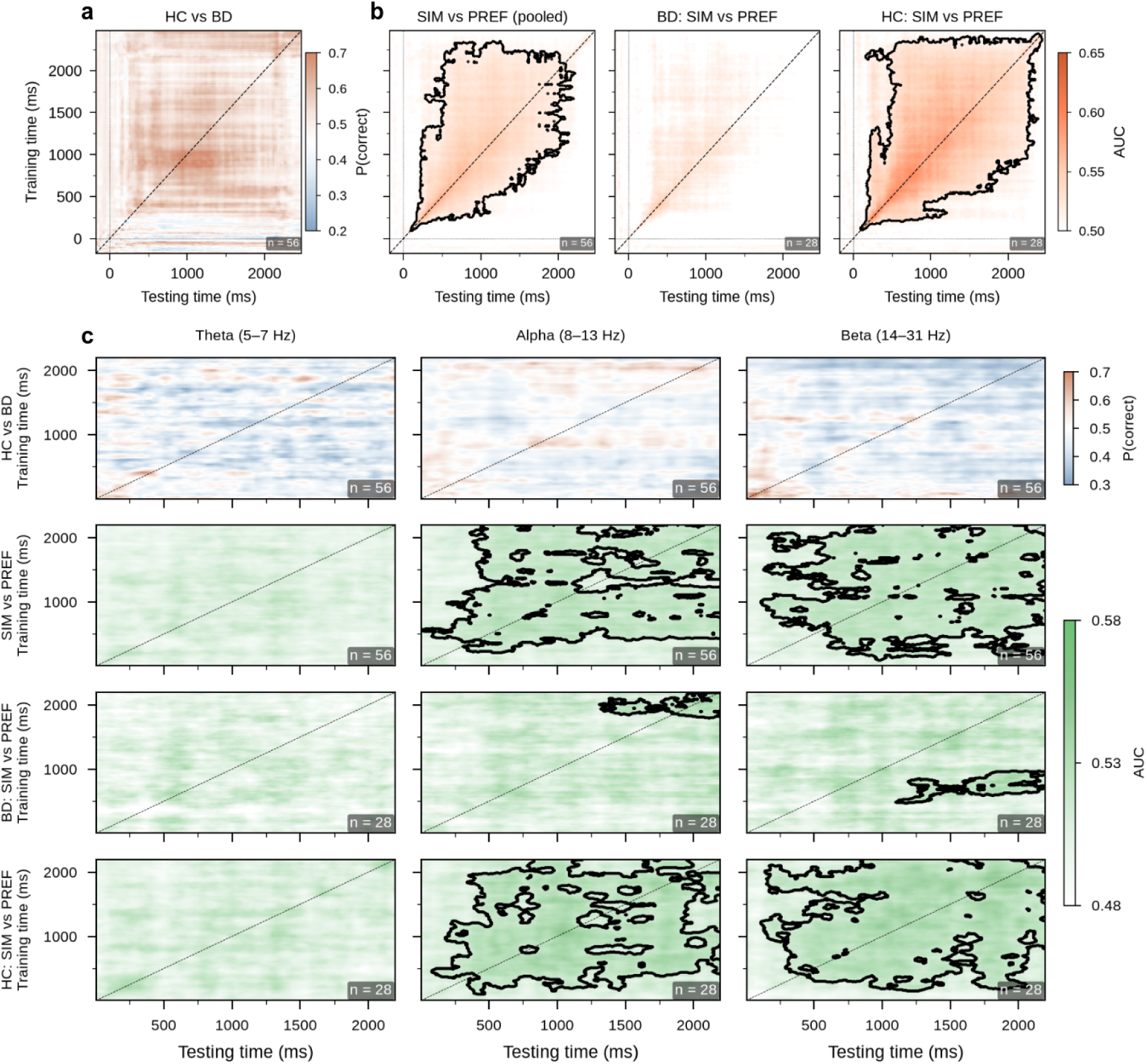
Whole-brain temporal generalisation decoding. Temporal generalisation matrices (train time × test time) for broadband and frequency band (theta, alpha, beta) data using all scalp channels. Each row shows four analyses: group decoding (HC vs BD, leave-one-out cross-validation), pooled condition decoding (SIM vs PREF), BD within-group decoding, and HC within-group decoding. Each cell represents classification performance (P(correct) for group decoding, AUC for condition decoding) of a linear discriminant analysis classifier. Significant clusters from two-dimensional cluster-based permutation tests (p < 0.05) are outlined. **(a)** No significant group effect. **(b)** Broadband (top row): significant condition effect, extensive temporal generalisation in HC (training 105 to 2435 ms, testing 95 to 2435 ms) but not in BD. **(c)** Theta (left column): no significant effects in any analysis. Alpha (middle column): significant condition effect, extensive temporal generalisation in HC (training 105 to 2195 ms, testing 85 to 2195 ms), marginal effect in BD (training 1795 to 2185 ms, testing 1315 to 2195 ms). Beta (right column): significant condition effect, extensive temporal generalisation in HC (training 155 to 2195 ms, testing 115 to 2195 ms), marginal effect in BD (training 475 to 965 ms, testing 1105 to 2195 ms). The dashed line indicates the diagonal. n = 56 for group and pooled condition; n = 28 per group for within-group analyses.

Frequency band analyses revealed a triple dissociation (Figure 2c). Theta produced no significant effects in any analysis (Figure 2c, left column). Alpha showed a significant condition effect (p < 0.001; Figure 2c, middle column), with extensive temporal generalisation in controls (p < 0.01) but only a marginal, late-emerging cluster in patients (p < 0.05). Beta similarly showed a significant condition effect (p < 0.001; Figure 2c, right column), with extensive temporal generalisation in controls (p < 0.001) and a marginal, restricted cluster in patients (p = 0.048). The direct group comparison did not reach significance across these three frequency bands (Figure 2c top row).

### Anterior region of interest temporal generalisation decoding

To further delineate the spatial distribution of these stable temporal codes, decoding was next restricted to an anterior region of interest. The anterior region of interest analysis revealed a striking reversal of the whole-brain theta pattern (Figure 3). Anterior theta showed a significant condition effect (p < 0.01; Figure 3f), and within-group analysis revealed significant temporal generalisation in patients (p < 0.01, training 325 to 1675 ms, testing 265 to 2025 ms; Figure 3j) but not in controls (Figure 3n). This indicates that anterior theta carries self versus non-self information specifically in patients. Anterior alpha showed significant temporal generalisation in controls (p < 0.05; Figure 3o) but not patients (Figure 3k). Anterior beta showed significant temporal generalisation in both groups (patients: p < 0.05 and p < 0.05, Figure 3l; controls: p < 0.001, Figure 3p).

**Figure 3.**
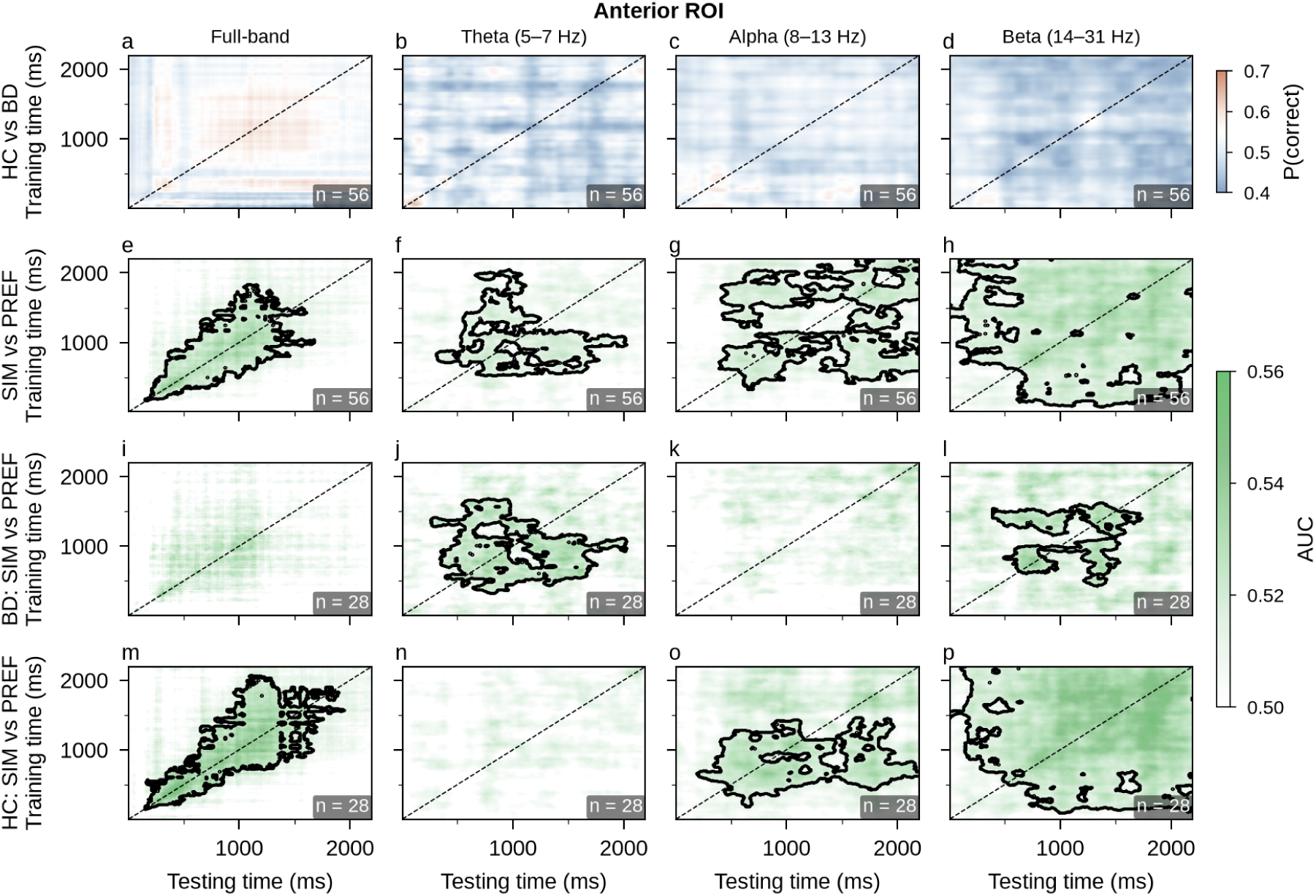
Anterior region of interest temporal generalisation decoding. Same format as Figure 2c but restricted to anterior electrodes (Fp1, Fp2, F7, F3, Fz, F4, F1, F2, F8). Full-band (first column): significant condition effect (p = 0.030), HC significant (p = 0.021), BD not significant. Theta (second column): significant condition effect (p = 0.005) and a reversal of the whole-brain pattern — BD showed significant temporal generalisation (p = 0.0026, training 325 to 1675 ms, testing 265 to 2025 ms) while HC did not, indicating that anterior theta carries self versus non-self information specifically in patients. Alpha (third column): significant condition effect (p = 0.005), HC significant (p = 0.017), BD not significant. Beta (fourth column): significant condition effect (p = 0.0002), with both groups showing significant within-group temporal generalisation (BD: p = 0.045 and p = 0.040; HC: p = 0.0002). n = 56 for group and pooled condition; n = 28 per group for within-group analyses.

### Posterior region of interest temporal generalisation decoding

Finally, to further delineate the spatial distribution of these neural codes, temporal generalisation decoding was evaluated within a posterior region of interest. Posterior region of interest analyses revealed more robust frequency band effects in patients than the whole-brain analysis indicated (Figure 4). Posterior alpha showed significant temporal generalisation in both controls (p < 0.01, Figure 4o) and patients (p < 0.05 and p < 0.05, Figure 4k), indicating partially preserved posterior alpha coding.

**Figure 4.**
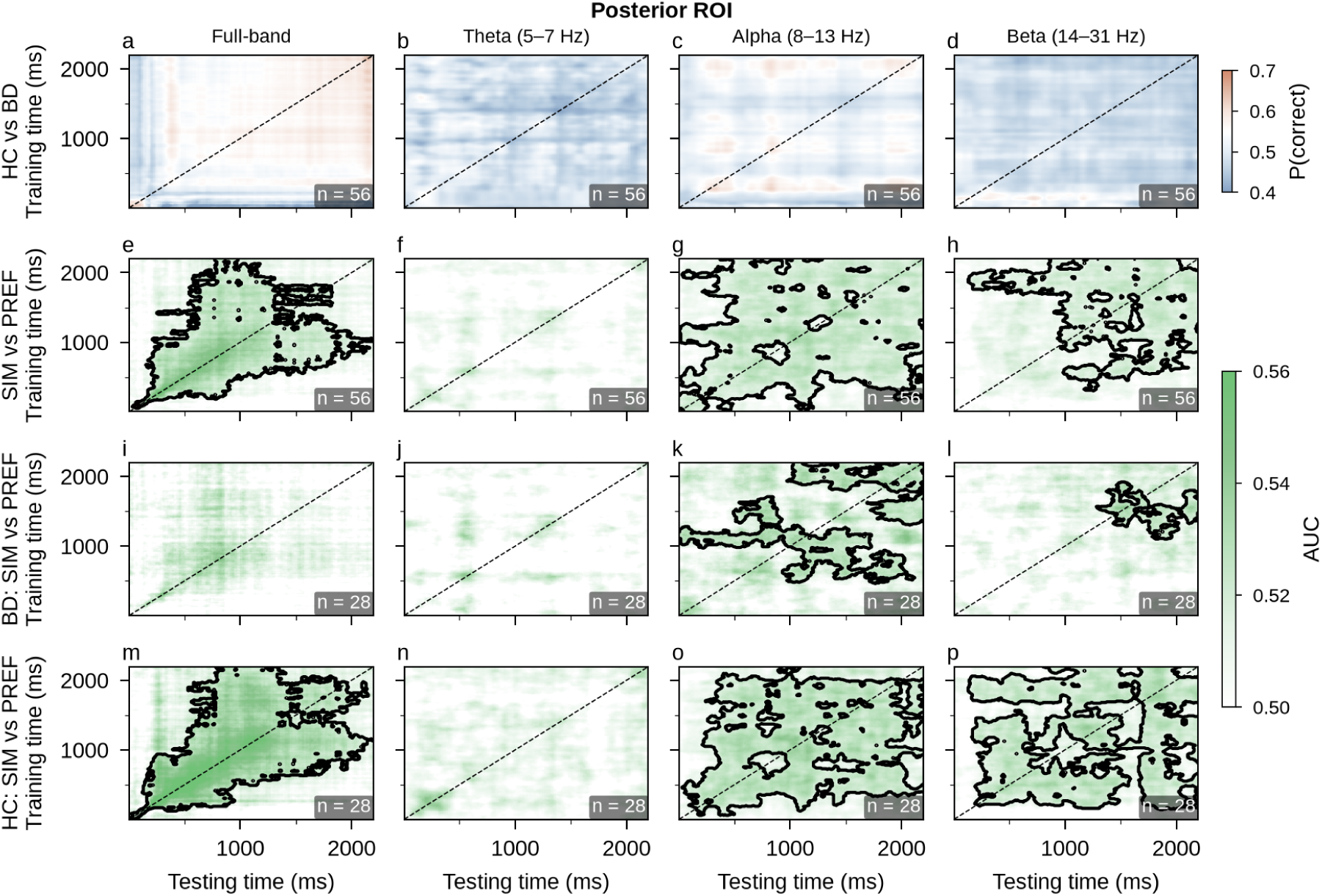
Posterior region of interest temporal generalisation decoding. Same format as Figure 3 but restricted to posterior electrodes (P3, Pz, P4, POz, O1, Oz, O2). Full-band (first column): significant condition effect (p = 0.007), HC significant (p = 0.007), BD not significant. Theta (second column): no significant condition or within-group effects. Alpha (third column): significant condition effect (p = 0.0006), with significant temporal generalisation in both HC (p = 0.0034) and BD (two clusters: p = 0.017 and p = 0.025), indicating more robust posterior alpha coding in BD at the regional level than in the whole-brain analysis. Beta (fourth column): significant condition effect (p = 0.002), with significant temporal generalisation in both HC (p = 0.001) and BD (p = 0.038). n = 56 for group and pooled condition; n = 28 per group for within-group analyses.

Posterior beta similarly showed significant effects in both groups (controls: p < 0.01, Figure 4p; patients: p < 0.05, Figure 4l). Posterior theta showed no significant condition or within-group effects (Figure 4b, 4f).

### Clinical correlations with anterior temporal generalisation

To determine whether the strength of these neural representations relates to psychological traits and symptom severity, we examined the relationship between decoding performance and clinical measures. Per-subject mean AUC values were extracted from significant anterior temporal generalisation clusters and correlated with clinical measures (Figure 5). These correlations were exploratory and uncorrected for multiple comparisons. Anterior theta condition decoding AUC correlated positively with RRS total scores across all participants (ρ = 0.322, p = 0.015, n = 56; Figure 5 right column). Anterior alpha condition decoding AUC correlated negatively with HAMA scores in patients (ρ = -0.434, p = 0.034, n = 24; Figure 5 the middle column), and anterior beta within-group decoding AUC in patients correlated negatively with RRS total scores (ρ = -0.391, p = 0.040, n = 28). Neither YMRS nor BPRS scores correlated with any decoding metric.

**Figure 5.**
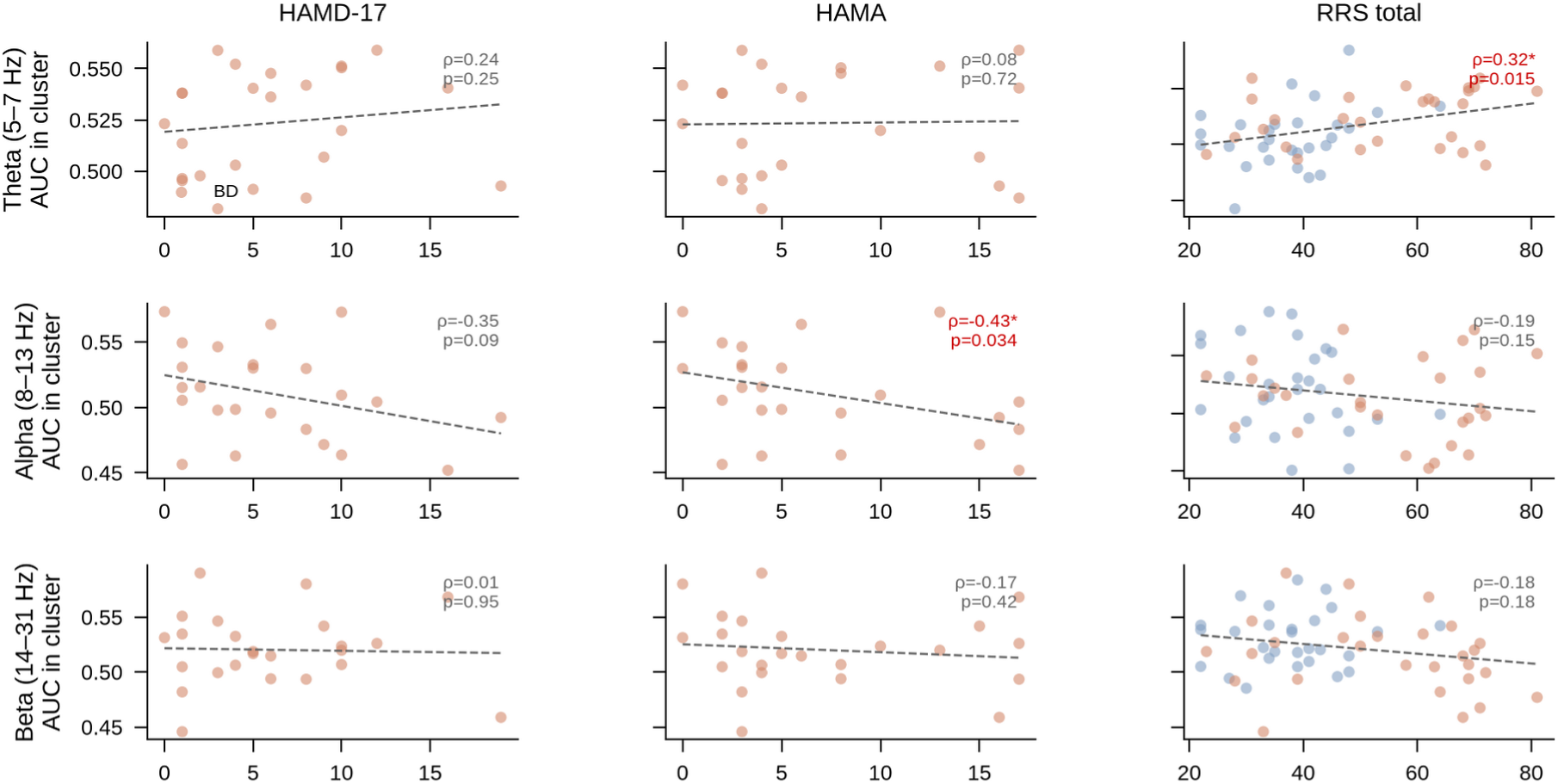
Clinical correlations with anterior temporal generalisation decoding. Scatter plots showing the relationship between per-subject mean AUC within significant anterior temporal generalisation clusters and clinical measures (HAMD-17, HAMA, and RRS total), for theta (top row), alpha (middle row), and beta (bottom row) frequency bands. AUC values were extracted from the condition decoding analysis (pooled across groups). Healthy controls (HC) are shown in blue and patients with bipolar disorder (BD) in orange. Dashed lines indicate linear fits. Spearman correlations and p values are displayed for each panel. Three significant correlations were observed: anterior theta condition AUC correlated positively with RRS total across all participants (ρ = 0.322, p = 0.015, n = 56), anterior alpha condition AUC correlated negatively with HAMA in BD (ρ = -0.434, p = 0.034, n = 24), and anterior beta BD-specific AUC correlated negatively with RRS total in BD (ρ = -0.391, p = 0.040, n = 28).

### Summary and Discussion

Using an emotionally neutral paradigm, we found that the difference in event-related potentials between self-related and non-self-related conditions was smaller in patients than controls. We then asked whether a response pattern that discriminates the two conditions at one moment also discriminates at later moments. In controls, the pattern separating preference from similarity trials early in the trial could be recovered throughout the remainder of the trial (King & Dehaene, 2014); in patients, no such generalisation emerged. Exploratory analyses further suggested that the oscillatory and spatial organisation of this difference varies between groups, though these observations require replication.

What this persistence reflects cannot be determined from the present design. Because responses were permitted only after the 2000 ms stimulus period, participants had to hold their decision across the trial, but what performance required was retention of the chosen side rather than of the evaluative mode that produced it. Prolonged condition coding in controls may therefore reflect a sustained task set, continued evaluative elaboration after the decision is reached, or another persistent state difference between conditions, none of which is necessary for a correct response. On this reading, the group difference would indicate that controls sustain a task-irrelevant state that patients do not, rather than that patients fail at something the task demands, consistent with both groups performing the task effectively. Response maintenance alone is an unlikely account, since both conditions required the same binary response and a response code would not discriminate between them. Because each condition was performed as a block, the sustained difference may also reflect the two task states themselves rather than trial-by-trial engagement of self-reference. Distinguishing these accounts would require varying the duration of the delay or probing the content of what is held.

That the event-related potential and decoding analyses did not converge on the same group differences has a straightforward explanation. The group effect in the event-related potentials was a main effect: amplitudes were lower in patients across both conditions, at 18 of 26 channels and over most of the epoch. Condition decoding was computed within each participant on the contrast between the two conditions, so any difference common to both cancels and cannot contribute to classification. More generally, univariate analyses are sensitive to variability between participants in mean response magnitude, whereas multivariate classifiers operate on the spatial pattern across channels after accounting for its covariance and are comparatively insensitive to it (Davis et al., 2014). Decoding also estimates an accuracy value for each participant before those values are tested at the group level, so between-participant variability in classification performance is added to whatever neural difference is present; at the sample sizes typical of clinical electroencephalography these estimates are correspondingly unstable, and effects detectable univariately may not be recovered by classification (Jamalabadi et al., 2016).

The frequency band, regional, and correlation analyses were exploratory. Alpha and beta carried self versus non-self information in both groups, consistent with established roles for alpha in gating information flow between cortical networks (Jensen & Mazaheri, 2010; Klimesch, 2012) and for beta in maintaining the current cognitive set (Engel & Fries, 2010) and both showed more temporally extensive generalisation in controls. Theta produced a different pattern: no generalisation at the whole-brain level in either group, but significant generalisation in patients at anterior sites, where controls showed none. Frontal midline theta, generated by medial prefrontal and anterior cingulate cortices, supports internally directed evaluation and conflict monitoring (Cavanagh & Frank, 2014; Knyazev, 2012), and was the oscillatory correlate of self-referential processing abnormalities in our previous study using the same paradigm (Hsu et al., 2021), raising the possibility that it contributes to self-referential processing differently in bipolar disorder than in healthy individuals.

The exploratory clinical correlations showed a consistent pattern worth noting. Anterior theta temporal generalization correlated positively with rumination across all participants, while anterior alpha correlated negatively with anxiety and anterior beta correlated negatively with rumination in patients. Notably, neither manic symptoms nor general psychopathology as indexed by the YMRS and BPRS correlated with any decoding metric. All associations that did emerge involved the depressive, anxious, and ruminative dimension — clinical features consistently linked to default mode network engagement and internally directed thought (Chou et al., 2023; Hamilton et al., 2015; Jamalabadi et al., 2016; Riaz et al., 2025) — which is the network engaged by the preference condition in this paradigm (Ratnasari et al., 2025). This convergence raises the possibility that residual internalizing symptoms, even at subclinical levels during clinical stability, shape how self-referential processing is neurally implemented. These correlations were uncorrected and based on modest sample sizes, so this interpretation remains tentative.

These results may be compared with those from a study using the same paradigm in patients with major depressive disorder (Hsu et al., 2021), where patients showed larger differences between conditions in frontocentral late positive potentials, accompanied by elevated theta power during the self-related condition, with the magnitude of the difference correlating positively with rumination. The direction of the amplitude effect in the present study was the opposite. Frontal theta was associated with rumination in both datasets, although the present result came from an exploratory regional analysis and the two theta measures are not equivalent. Task modifications between the studies and separate recruitment make the comparison indirect. With those caveats, the observation that condition differences were larger in depression and smaller in bipolar disorder, using stimuli free of emotional valence, is compatible with self-related processing being altered in both conditions in different ways. Whether this pattern relates to perseverative self-focused thought in depression (Nolen-Hoeksema et al., 2008) versus unstable self-evaluation in bipolar disorder (Inder et al., 2008) is a question these data cannot address; a single study including both diagnostic groups, ideally with measures of subjective cognitive function alongside objective assessment (Miskowiak et al., 2018), and application of the task during bipolar depression, would be informative.

Several limitations should be noted. The decoding analyses did not include a test of the group by condition interaction, so the contrast between significant temporal generalisation in controls and its absence in patients should not be read as a demonstrated group difference. All patients were taking psychotropic medication, which can alter oscillatory activity. The frequency band analyses used oscillatory power, which is insensitive to phase information. Some patients had symptom scores above conventional euthymia thresholds, and the cross-sectional design cannot establish whether the observed differences represent stable trait features. The sample was predominantly female and consisted entirely of bipolar I patients, and the sample size (28 per group), while comparable to previous temporal generalisation studies in clinical populations (Jeong et al., 2026) and healthy participants (Xie et al., 2020), limits statistical power.

In conclusion, the difference between self-related and non-self-related judgements was smaller in patients with bipolar disorder than in controls, and only in controls did the neural pattern separating the two conditions remain decodable across the trial. This distinction between the size of a condition difference and its persistence is not available from conventional amplitude analyses, and raises the question of whether it relates to the self-related difficulties reported in this population. Exploratory analyses pointing to altered frontal theta engagement, and to associations with residual depressive, anxious, and ruminative symptoms, indicate where a mechanism might be sought, though they require replication. More broadly, these findings show what temporal decoding can add to the characterisation of cognition in bipolar disorder.

## Data Availability

All data produced in the present study are available upon reasonable request to the authors.

## Acknowledgements

The authors thank all participants for their time and effort. This work was supported by the National Science and Technology Council (113-2410-H-008-082, 114-2410- H-008-066, 115-2410-H-008-051-MY3) and the Taiwan Ministry of Education Higher Education Sprout Project to TYH; by the National Science and Technology Council (113-2423-H-038-002-MY3, 114-2410-H-038-037) to NWD; and by Taipei Medical University and Taipei Medical University Hospital (112TMU-TMUH-09) to PHC.

## Author contributions

**Pao-Huan Chen:** Conceptualisation, Funding, Investigation, Data Curation, Writing- Original draft preparation, Supervision

**Niall W. Duncan**: Conceptualisation, Funding, Writing- Original draft preparation, Supervision

**Yi-Ju Liu**: Investigation, Software

**Hsin-Chien Lee**: Investigation, Data Curation, Writing- Original draft preparation

**Tzu-Yu Hsu**: Conceptualisation, Funding, Investigation, Methodology, Data Curation, Software, Visualisation, Writing- Original draft preparation, Supervision

## Conflicts of interest

The authors declare no conflicts of interest.

## Declaration of generative AI in the writing process

During the preparation of this work the authors used Claude (Anthropic) to assist with drafting and revising the manuscript and with literature searching. All analyses were designed and carried out by the authors. After using this tool the authors reviewed and edited the content, verified all cited references against the primary sources, and take full responsibility for the content of the publication.

---

**Supplementary table 1.** Demographic and clinical characteristics of the study sample.

|  | HC (n = 34) | BD (n = 30) | Statistic | p |
| --- | --- | --- | --- | --- |
| Age (years) | 31.9 ± 7.2 | 30.6 ± 8.0 | t = 0.66 | 0.51 |
| Sex (M/F) | 9/25 | 8/22 | $\chi^2 = 0.00$ | 1 |
| Education (years) | 16.5 ± 1.6 | 15.1 ± 2.0 | t = 3.06 | <.01 |
| RRS total | 38.0 ± 10.1 | 52.9 ± 16.7 | t = -4.38 | <.01 |
| YMRS |  | 1.0 ± 2.7 |  |  |
| HAMD-17 |  | 6.0 ± 4.9 |  |  |
| HAMA |  | 6.7 ± 5.6 |  |  |
| BPRS |  | 25.6 ± 6.1 |  |  |
| Mood stabilizers (n) |  | 27 (90%) |  |  |
| Lithium |  | 0 (0%) |  |  |
| Lamotrigine |  | 8 (29%) |  |  |
| Valproate |  | 4 (13%) |  |  |
| Antipsychotics (n) |  | 24 (80%) |  |  |
| Antidepressants (n) |  | 13 (43%) |  |  |
| Benzodiazepines (n) |  | 15 (50%) |  |  |
| **Total medications** |  | 3.0 ± 1.0 |  |  |

Note. Data are presented as mean ± standard deviation or frequencies. HC = Healthy Controls; BD = Bipolar Disorder; RRS = Ruminative Responses Scale; YMRS = Young Mania Rating Scale.

**Supplementary table 2.** Demographic and clinical characteristics of the study sample in EEG analysis.

|  | HC<br>(n = 28) | BD<br>(n = 28) | Statistic | p |
| --- | --- | --- | --- | --- |
| Age (years) | 31.2 ± 6.9 | 30.6 ± 7.9 | t = 0.34 | 0.73 |
| Sex (M/F) | 6/22 | 7/21 | $\chi^2 = 0.00$ | 1 |
| Education (years) | 16.2 ± 0.6 | 15.1 ± 2.1 | t = 2.77 | <.01 |
| RRS total | 37.8 ± 9.7 | 54.2 ± 16.5 | t = -4.55 | <.01 |
| YMRS |  | 0.5 ± 1.7 |  |  |
| HAMD-17 |  | 6.0 ± 5.0 |  |  |
| HAMA |  | 7.0 ± 5.8 |  |  |
| BPRS |  | 25.4 ± 6.3 |  |  |
| Mood stabilizers (n) |  | 25 (89%) |  |  |
| Lithium |  | 0 (0%) |  |  |
| Lamotrigine |  | 8 (29%) |  |  |
| Valproate |  | 3 (11%) |  |  |
| Antipsychotics (n) |  | 22 (79%) |  |  |
| Antidepressants (n) |  | 12 (43%) |  |  |
| Benzodiazepines (n) |  | 14 (50%) |  |  |
| **Total medications** |  | 3.0 ± 1.0 |  |  |

Note. Data are presented as mean ± standard deviation or frequencies. HC = Healthy Controls; BD = Bipolar Disorder; RRS = Ruminative Responses Scale; YMRS = Young Mania Rating Scale.

## SUPPLEMENTARY METHODS

### Participants

Exclusion criteria for patients included comorbid neurological disorders, substance use disorders, and poor visual acuity. Healthy controls were recruited through advertisements on the public websites of the affiliated hospitals and university.

Of the 34 controls and 32 patients initially recruited, two patients did not complete the colour task, leaving a behavioural analysis sample of 64 participants (34 controls, 30 patients). Prior to electroencephalographic analysis, eight further participants (six controls, two patients) were excluded for falling below the minimum threshold of 20 valid trials per condition after artifact rejection, giving a final electroencephalographic sample of 28 controls and 28 patients.

### Colour judgment task

The task was adapted from the colour judgment paradigm used in our previous study of major depressive disorder (Hsu et al., 2021; Johnson et al., 2005). Colours were selected from a set of 12 equiluminant values spaced at 30° intervals around the a*(red–green) and b*(yellow–blue) plane of CIELAB colour space. In the similarity condition, the two comparison squares differed in distance from the target by one and two colour steps (30° and 60°). In the preference condition, both comparison squares were two steps (60°) from the target.

Participants were informed which condition they would perform before each block, and breaks were provided every 50 trials. The task was programmed and presented using the Psychophysics Toolbox (Brainard, 1997; Kleiner et al., 2007; Pelli, 1997) running in MATLAB.

Two modifications were made relative to Hsu et al. (2021): the difficulty manipulation was removed in favour of a fixed colour distance, and the preference condition used equidistant comparison colours to eliminate any implicit correct answer.

### Electroencephalographic recording and preprocessing

Recording used Ag/AgCl electrodes mounted in an elastic cap (Easycap; Brain Products GmbH) in a 30-electrode arrangement following the international 10–20 system. Six additional channels recorded vertical and horizontal electrooculogram and mastoid references. Online recording used BrainAmp with a bandpass filter of 0.05 to 1000 Hz and a sampling rate of 1000 Hz.

Continuous data were resampled to 200 Hz and filtered at 1 to 30 Hz before independent component analysis, which was performed using the FastICA algorithm with 15 components to identify signals arising from eye movements, blinks, and muscle activity. Cleaned data were then filtered at 0.1 to 45 Hz. Missing channels were reconstructed using spherical spline interpolation.

For event-related potential analyses, similarity trials with no response or incorrect responses were excluded, and preference trials with no response were excluded. Data were segmented from −200 to 2500 ms relative to stimulus onset and baseline-corrected using the −200 to 0 ms prestimulus interval, which removes slow voltage drifts affecting amplitude comparisons across conditions. Epochs exceeding ±100 µV in any channel were rejected.

For frequency band analyses, the same trial exclusion criteria were applied, but data were segmented from −1000 to 2500 ms without time-domain baseline correction, because the subsequent Morlet wavelet decomposition requires preservation of low-frequency oscillatory information that baseline subtraction would distort. Baseline normalisation was instead applied in the frequency domain. The same ±100 µV rejection criterion was used. Following artifact rejection and behavioural trial filtering, trial counts were equalised across conditions within each participant by random subsampling.

### Event-related potential analysis

Event-related potentials were computed by averaging artifact-free, valid trials within each condition for each participant. Group, condition, and group by condition interaction effects were tested using spatiotemporal cluster-based permutation tests (5000 permutations) across all electroencephalographic channels and the full epoch. Cluster-forming thresholds were set at p < 0.05 and clusters were considered significant at p < 0.05.

### Temporal generalisation decoding

Decoding was performed using MNE-Python (Gramfort et al., 2013) (Gramfort et al., 2013) with the GeneralizingEstimator framework (King et al., 2014; King & Dehaene, 2014). Classifiers trained at each time point were tested at all other time points, yielding a two-dimensional train-time by test-time matrix of decoding accuracy expressed as area under the receiver operating characteristic curve. A linear discriminant analysis classifier was trained with 5-fold stratified cross-validation to distinguish preference from similarity trials within each participant. Prior to decoding, epochs were smoothed using a 50 ms moving average with a 10 ms step, giving a temporal resolution of approximately 10 ms. Statistical significance was assessed using two-dimensional cluster-based permutation tests with spatial adjacency on the train-by-test grid (5000 permutations, cluster-forming threshold: one-sample t at p < 0.05, one-tailed) comparing accuracy against chance (0.5).

### Isolated frequency band decoding

No-baseline epochs were convolved with complex Morlet wavelets at 20 logarithmically spaced frequencies from 5 to 31 Hz using a constant wavelet length of 600 ms (number of cycles = frequency × 0.6), following Xie et al. (2020). Absolute power was computed and baseline-normalised in decibels using the −500 to −300 ms prestimulus interval. Power values were aggregated into three bands by averaging across frequency bins: theta (5 bins), alpha (6 bins), and beta (9 bins). Data were smoothed using a 50 ms moving average with a 10 ms step and cropped to 0 to 2200 ms before decoding. Decoding and statistical testing used the same classifier, cross-validation scheme, and permutation procedure as the broadband analysis.

### Region of interest decoding

The whole-brain analyses were repeated separately for anterior and posterior electrode subsets. The anterior region comprised Fp1, Fp2, F7, F3, Fz, F4, F1, F2, and F8; the posterior region comprised P3, Pz, P4, POz, O1, Oz, and O2. This division was motivated by the frontocentral topography of the late positive potential and the established association between frontal theta and self-referential processing (Hsu et al., 2021; Knyazev, 2013). Broadband, theta, alpha, and beta decoding was performed for each region using the same procedures, classifiers, and statistical testing as the whole-brain analyses.

### Clinical correlations

Per-participant mean AUC values were extracted from the significant cluster pixels identified in the anterior region of interest analyses and correlated with HAMD-17, HAMA, and RRS total scores using Spearman rank correlations. Correlations were computed within each group separately and, for the pooled condition analysis, across all participants. YMRS and BPRS were not included because the majority of patients had scores at or near floor on these scales.

### Statistical analysis

Behavioural data were analysed using independent-samples t-tests for accuracy and a 2 (condition: similarity, preference) × 2 (group: controls, patients) mixed analysis of variance for reaction times. Effect sizes are reported as Cohen’s d for t-tests and partial eta-squared for analysis of variance effects. Given the exploratory nature of the frequency band and region of interest analyses, and the absence of correction for the number of spectral and spatial comparisons performed, these findings are interpreted as hypothesis-generating. All analyses were performed using MNE-Python (Gramfort et al., 2013), SciPy, and pingouin.

## Notes

### Competing Interest Statement

The authors have declared no competing interest.

### Author Declarations

The study was approved by the Joint Institutional Review Board of Taipei Medical University.

